# Time-Dependent Variation in Immunoparalysis Biomarkers Among Patients with Sepsis and Critical Illness

**DOI:** 10.1101/2024.07.11.24310285

**Authors:** Abigail Samuelsen, Erik Lehman, Parker Burrows, Anthony S Bonavia

## Abstract

Immunoparalysis is a significant concern in patients with sepsis and critical illness, potentially leading to increased risk of secondary infections. This study aimed to perform a longitudinal assessment of immune function over the initial two weeks following the onset of sepsis and critical illness. We compared ex vivo stimulated cytokine release to traditional markers of immunoparalysis, including monocyte Human Leukocyte Antigen (mHLA)-DR expression and absolute lymphocyte count (ALC). A total of 64 critically ill patients were recruited in a tertiary care academic medical setting, including 31 septic and 33 non-septic patients. Results showed that while mHLA-DR expression significantly increased over time, this was primarily driven by the non-septic subset of critically ill patients. ALC recovery was more prominent in septic patients. Ex vivo stimulation revealed significant increases in TNF and IL-6 production over time in septic patients. However, IFNγ production varied with the stimulant used and did not show significant recovery when normalized to cell count. No significant correlation was found between mHLA-DR expression and other immunoparalysis biomarkers. These findings suggest the need for more nuanced immune monitoring approaches beyond the traditional ‘sepsis’ versus ‘non-sepsis’ classifications in critically ill patients. It also provided further evidence of a potential window for targeted immunotherapeutic interventions in the first week of critical illness.

## 1. Introduction

Sepsis, marked by host immune dysregulation and subsequent organ dysfunction, is a leading cause of critical illness, often resulting in death within days to months after the onset of acute illness [1]. Patient mortality rates in the intensive care unit have been cited as high as 30% [2-5]. Furthermore, secondary infection, resulting from sepsis-induced impairment of host immunity, is a well-recognized cause of morbidity [6]. One study found 69% of hospital readmissions were related to infection, with over 50% of those as recurrent or unresolved infections [7]. This ‘immunoparalysis’ is multifactorial [8,9], driven by immune cell exhaustion and apoptosis [10-12], anti-inflammatory cytokine production [13-16], metabolic dysfunction [17,18], and the expansion of regulatory T cells [19-22] and myeloid-derived suppressor cells [23,24].

Significant efforts have been made to identify immune phenotypes in sepsis to develop clinically useful risk-stratification tools that could improve clinical outcomes. A recent, landmark study by Venet and the REALISM (REAnimation Low Immune Status Marker) investigators identified a subgroup of severely injured patients who developed delayed injury-acquired immunodeficiency, independent of the primary disease [25]. These findings suggest that immunoparalysis is not specific to sepsis but can be assessed in critically ill patients by monitoring a common panel of complementary markers of pro-/anti-inflammatory, innate, and adaptive immune responses. Moreover, this research highlights the urgent need to identify routinely accessible immunosurveillance markers to pinpoint patients who might benefit from customized immunoadjuvant therapies [26]. Given the rapid progression of sepsis and the narrow window for effective clinical intervention, it is also important that these biomarkers can be rapidly processed in a point-of-care clinical setting.

Based in part on these insights, our lab has pioneered a fast and precise assay for detecting immunoparalysis. We recently reported that ex vivo stimulation of both innate and adaptive immune system components did not predict secondary infection rates but reliably predicted organ function within 48 hours of the assay [27]. Clustering analysis further revealed two distinct immune phenotypes, characterized by differential responses to 18 hours of lipopolysaccharide (LPS, or endotoxin) and 4 hours of anti-CD3/anti-CD28 stimulation.

The present analysis aimed to use these assays to survey immune function over the first two weeks following the onset of sepsis and critical illness. Specifically, our goal was to compare ex vivo, stimulated cytokine release to traditional markers of immunoparalysis, including monocyte Human Leukocyte Antigen (mHLA)-DR expression (a biomarker of innate immunity) and absolute lymphocyte count (ALC, a biomarker of adaptive immunity) [28]. We hypothesized that patterns in ex vivo, stimulated cytokine responses over time would mirror those of mHLA-DR expression and ALC. Due to challenges in reliably identifying secondary infections, including inconsistent diagnostic criteria and underreporting [29-31], we defined immunoparalysis based on immune parameters alone.

## 2. Results

### 2.1. Study Population

We recruited a total of 64 critically ill patients, including 31 individuals diagnosed with sepsis and 33 non-septic control (CINS) patients. A detailed breakdown of the demographic characteristics and clinical outcomes for these patients is provided in **Table 1**. For septic patients with confirmed infections, the specific microbial sources are listed in **Appendix Table A1**. The characteristics of the CINS control group are comprehensively detailed in **Appendix Table A2**. Notably, during the 30-day follow-up period, two septic patients and two CINS patients were lost to follow-up, as illustrated in **Appendix Figure B1**.

**Table 1.** Patient Demographics and Clinical Outcomes. ^*^ within 14 days following enrollment. P-values represent two-sample t-test comparing group means for normally distributed variables, Wilcoxon Rank Sum test comparing group medians for skewed variables, or Chi-square test comparing proportions for categorical variables.

2.1 Study Population
| Demographic and Clinical Characteristics | Critically Ill, Septic<br>(n = 31) | Critically Ill, Non-Septic<br>(n = 33) | P-value |
| --- | --- | --- | --- |
| Age (mean ± SD) | 65.5 ± 13.4 | 64.1 ± 15.4 | 0.708 |
| Female (n, %) | 14 (48) | 15 (52) | 0.981 |
| **Infection etiology (n, %)** |  |  |  |
| ---Gram-negative | 11 | N/A |  |
| ---Gram-positive | 9 | N/A |  |
| ---Mixed | 11 | N/A |  |
| ---Clinical diagnosis only, no positive cultures | 2 | N/A |  |
| **Severity of illness** |  |  |  |
| APACHE II Score (mean ± SD) | 21.1 ± 7.1 | 18.4 ± 6.7 | 0.123 |
| SOFA Score (median, IQR) | 8.0 (7.1, 9.7) | 6.0 (5.7, 7.9) | 0.042 |
| Charlson Comorbidity Index (median, IQR) | 5.0 (4.0, 5.9) | 4.0 (3.0, 4.9) | 0.113 |
| Patients receiving stress-dosed hydrocortisone ≥ 8 h (n, %) | 9 (29.0) | 5 (15.2) | 0.179 |
| Daily hydrocortisone dose (mg) (median, IQR) | 150 (119, 188) | 200 (129, 264) | 0.202 |
| Duration of hydrocortisone (days*) (median, IQR) | 3.0 (1.9, 3.9) | 2.0 (0.6, 3.8) | 0.374 |
| **Laboratory Values** |  |  |  |
| Leukocyte Count, $\times 10^3/\mu\text{l}$ (median, IQR) | 14.5 (13.4, 20.7) | 10.8 (10.7, 14.8) | 0.055 |
| Absolute Lymphocyte Count, $\times 10^3/\mu\text{l}$ (median, IQR) | 0.66 (0.54, 0.78) | 1.01 (0.87, 1.54) | < 0.001 |
| Absolute Monocyte Count, $\times 10^3/\mu\text{l}$ (median, IQR) | 0.47 (0.34, 0.68) | 0.78 (0.74, 1.15) | < 0.001 |
| Lactic Acid (mg/dL) on admission (median, IQR) | 3.8 (3.2, 4.9) | 2.7 (2.2, 4.2) | 0.112 |
| Shock (vasopressors and lactate ≥ 2) on admission (n, %) | 10 (32.3) | 10 (30.3) | 0.866 |
| **Short-Term Outcomes** |  |  |  |
| Proportion of Cohort developing Secondary Infections (n, %) | 1 (3.2) | 2 (6.1) | 1.000 |
| 30-Day Mortality Rate (n, %) | 3 (4.7) | 6 (9.4) | 0.478 |
| Hospital Length of Stay, days (median, IQR) | 10.0 (9.5, 17.4) | 8.0 (8.3, 21.0) | 0.472 |
| 30-Day Hospital Readmission amongst survivors (n<sub>alive</sub>/n<sub>30-day survivors</sub>, %) | 4/28 (14.3) | 2/25 (8.0) | 0.423 |
| Discharged status |  |  |  |
| ---Home (n, %) | 13 (22.0) | 15 (25.4) | 0.880 |
| ---Skilled nursing facility (n, %) | 13 (22.0) | 11 (18.6) | 0.393 |
| ---Long-term acute care hospital (n, %) | 2 (3.4) | 1 (1.7) | 0.592 |
| ---Hospice (n, %) | 0 | 1 (1.7) | 1.000 |

### 2.2. mHLA-DR Expression Does Not Differ Between Critically Ill Patients With and Without Sepsis

While mHLA-DR expression significantly increases over the first 14 days following onset of critical illness (p = 0.002), this increase is driven primarily by non-septic patients (p = 0.006), with septic patients exhibiting only a trend towards significance over this time (p = 0.057, **Fig 1**). There was no significant difference between the slopes (i.e., rates of immune recovery, as assessed by mHLA-DR expression over the 14-day period) of critically ill patients with or without sepsis (p = 0.688).

**Figure 1.**
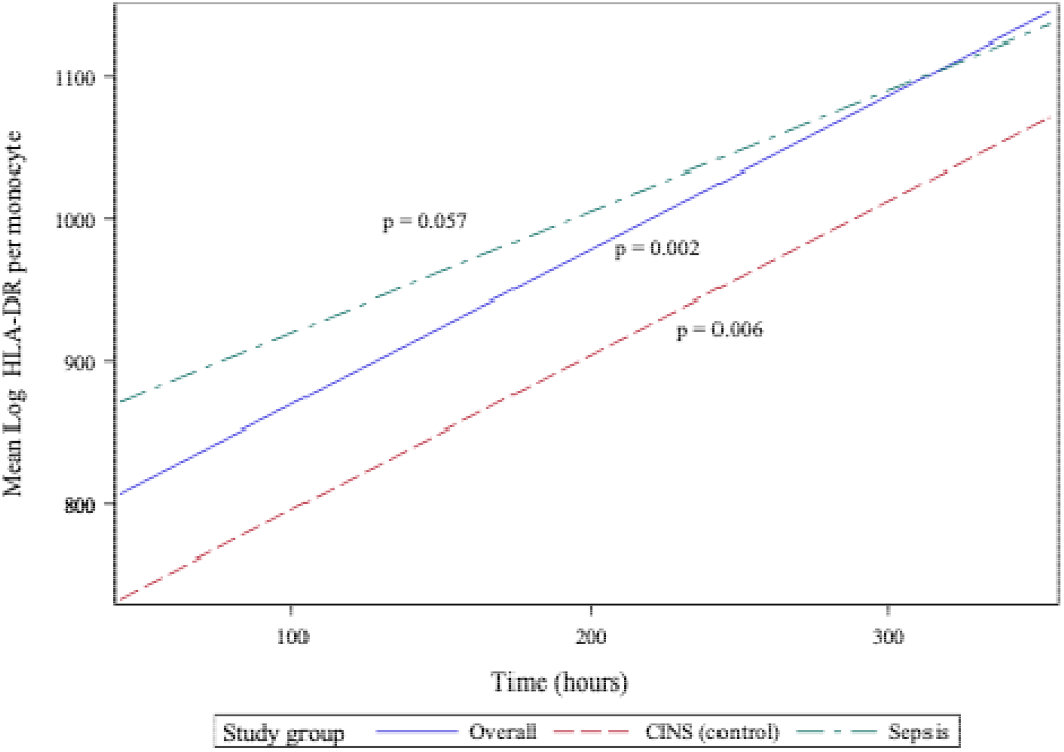
Monocyte Human Leukocyte Antigen (mHLA)-DR expression over the initial 14 days following critical illness. *P*-values indicate the significance of the rate of change in mHLA-DR expression in each subgroup over time, as analyzed using a mixed-effects linear regression model.

### 2.3. Absolute Lymphocyte Count Does Not Differ Between Critically Ill Patients With and Without Sepsis

Absolute lymphocyte count (ALC) also significantly recovers over the 14 days following onset of critical illness (p < 0.001, 95% CI 0.072 - 0.259, **Fig 2**). However, in contrast to the pattern of mHLA-DR expression, this recovery is primarily driven by the septic subset of critically ill patients (p = 0.003, 95% CI 0.073 - 0.332), with the CINS subgroup exhibiting a trend towards significance over this period (p = 0.059). Although the rates of ALC recovery do not vary significantly between subgroups (p = 0.436), there are notable differences between the mean ALC values of each sub-group on days 1 and 7 (p < 0.001 and p = 0.006, respectively). By day 14, these differences are no longer significant (p = 0.195).

**Figure 2.**
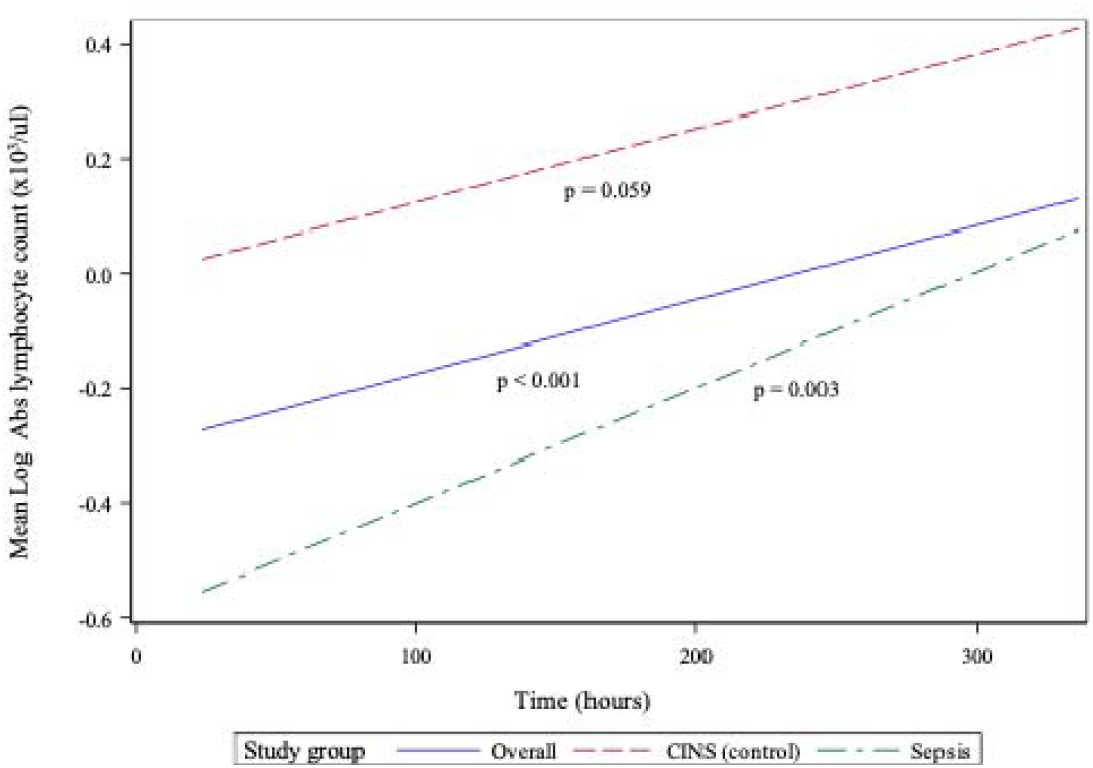
Absolute lymphocyte count (ALC) over initial 14 days following critical illness. *P*-values indicate the significance of the rate of change in ALC in each subgroup over time, as analyzed using a mixed-effects linear regression model.

### 2.4. Tumor Necrosis Factor Production Following Ex Vivo Endotoxin Stimulation of Whole Blood Does Not Differ Between Critically Ill Patients with and without Sepsis

LPS-stimulated cytokine production recapitulates the pattern of immune recovery modeled by mHLA-DR and ALC (**Fig 3A**). Specifically, following 18 hours of endotoxin stimulation, TNF production was observed to recover significantly over time in the combined cohort (p < 0.001, 95% CI 0.115 - 0.398). TNF production in septic patients increased significantly over the 14-day observation period (p < 0.001, 95% CI 0.195 - 0.615), although not in the CINS subset (p = 0.253). Rates of TNF recovery differed between subgroups (p = 0.041), driven primarily by differences in the concentration of this cytokine on days 1 (p < 0.001) and 7 (p = 0.001) of whole blood stimulation.

**Figure 3.**
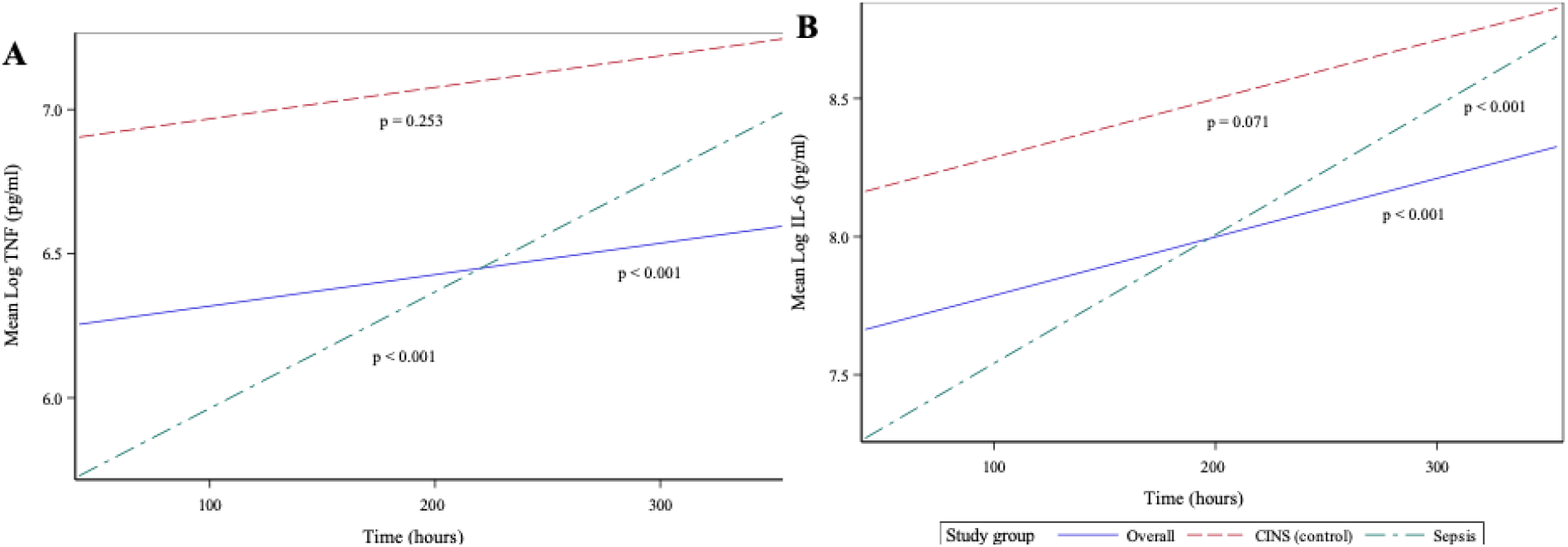
Ex vivo production of cytokines over time, following 18 hours of endotoxin stimulation of whole blood sampled from critically ill patients. (A) Stimulated tumor necrosis factor (TNF) production, (B) Stimulated interleukin (IL)-6 production. *P*-values indicate the significance of the rate of change in cytokine concentrations in each subgroup over time, as analyzed using a mixed-effects linear regression model.

The pattern of LPS-stimulated interleukin (IL)-6 was closely related to TNF production. In critically ill patients, IL-6 significantly increased over time (p < 0.001, 95% CI 0.166 – 0.513, Fig 3B), primarily driven by the septic subgroup (p < 0.001, 95% CI 0.209 – 0.726) and with CINS patients demonstrating a trend towards significant IL-6 recovery over time (p = 0.071). Mean IL-6 production was significantly different between each subgroup, at 1- and 7-days post-sepsis (p = 0.002 and 0.038), but the overall rate of IL-6 recovery (i.e., slope of the concentration of time) over the 14-day period was not significantly different between subgroups (p = 0.147).

### 2.5. Interferon-gamma Response, following Ex Vivo stimulation of Whole Blood, Varies by Stimulant Used

T lymphocyte responses play a crucial role in the adaptive immune response to sepsis. To assess these responses, we stimulated whole blood using anti-CD3 and anti-CD28 antibodies for 18 hours. While this stimulation did not result in a significant overall difference in IFNγ production from days 1 to 14 following the onset of critical illness, we did observe specific differences between mean IFNγ concentrations in each subgroup. These differences were limited to days 1 (p = 0.048) and 14 (p = 0.043) of critical illness.

Non-specific T lymphocyte stimulation, using PMA, demonstrated a more dramatic change in IFNγ production over time. Specifically, cytokine production increased substantially over 14 days of critical illness in all patients (p < 0.001, 95% CI 0.116 – 0.407) and in the septic subset (p = 0.003, 95% CI 0.12 – 0.552). The CINS subset demonstrated a less dramatic change in PMA-induced IFNγ concentrations over this time (p = 0.059, 95% CI -0.008– 0.383). As with anti-CD3/anti-CD28 stimulation, mean IFNγ concentrations were most significant on days 1 (p = 0.006) and 7 (p = 0.030) following critical illness onset.

Since, lymphopenia is a critical component of sepsis and critical illness [31,32], we normalized interferon IFNγ production to cell count, thus distinguishing between quantitative and qualitative lymphocyte defects. When normalized to ALC, there was no statistically significant increase in stimulated IFNγ production over time. This held true whether T lymphocytes were stimulated using specific (anti-CD3/anti-CD28, **Fig 4A**) or non-specific (PMA, **Fig 4B)** agents. It also implies that, during critical illness, there is quantitative recovery of lymphocyte counts (**Fig 2**) although no qualitative recovery.

**Figure 4.**
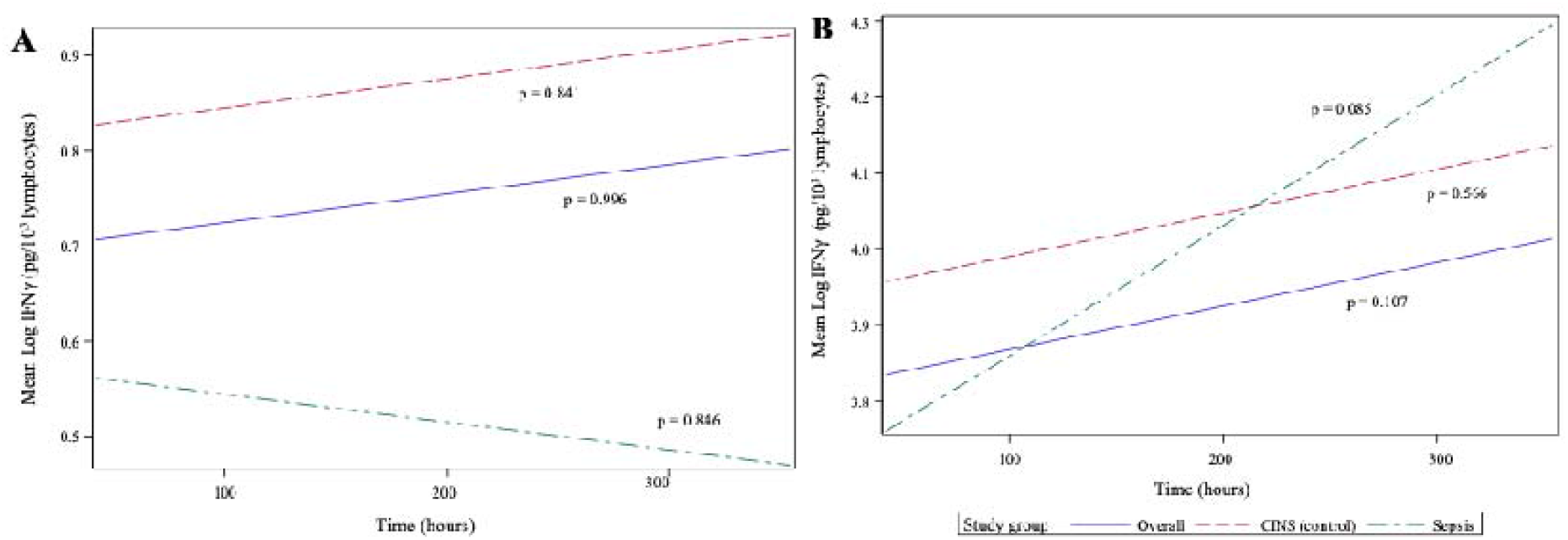
Ex vivo production of interferon (IFN)γover time, following 18 hours of (A) anti-CD3/anti-CD28 antibody, and (B) PMA. IFNγ concentrations have been normalized to lymphocyte count. P-values indicate the significance of the rate of change in IFNγ concentrations in each subgroup over time, as analyzed using a mixed-effects linear regression model.

### 2.6. mHLA-DR Is Not Correlated with Other Measured Markers of Immunoparalysis

Using repeated measures correlation (r_rm_), we examined the relationships among various markers of immunoparalysis. These included: (1) mHLA-DR expression, (2) ALC, (3) stimulated cytokine production, and (4) stimulated cytokine production normalized to lymphocyte count (in the case of IFNγ) and monocyte count (in the case of TNF).

Monocyte HLA-DR expression was not significantly associated with other immunoparalysis biomarkers. ALC demonstrated a weak correlation with endotoxin-stimulated TNF (r_rm_ = 0.336) and IL-6 (r_rm_ = 0.351). Additionally, ALC was weakly correlated with IFNγ concentration following anti-CD3/anti-CD28 (r_rm_ = 0.385) and PMA (r_rm_ = 0.347) stimulation.

Endotoxin-stimulated cytokines TNF and IL-6 were moderately correlated (r_rm_ = 0.495), with a higher correlation observed within the CINS subset (r_rm_ = 0.576). IFNγ concentration following T cell-specific (anti-CD3/anti-CD28) and non-specific (PMA) stimulation was also moderately correlated (r_rm_ = 0.446 in all patients, and 0.523 in CINS subset). When normalized to lymphocyte count, these responses to T cell stimulants remained moderately correlated (r_rm_ = 0.359 in all patients, and 0.529 in CINS subset).

### 2.7. While Biomarkers of Immunoparalysis Are Not Associated with Secondary Infection Rate, Endotoxin-Associated IL-6 Production Is Associated with 30-Day Hospital Readmission

There was no association between the concentration of each biomarker and 30-day mortality or the number of days from critical illness until death. Neither did we discover any association with 30-day secondary infection rates. In the analysis of hospital length of stay following critical illness with or without sepsis diagnosis, we found statistically significant intercepts in several quantile regression models, though these were not accompanied by associations with any of the independent variables. These findings may reflect an intrinsic difference in hospital length of stay between sepsis and CINS patients, not explained by the biomarkers used in our models.

Logistic regression revealed a statistically significant relationship between endotoxin-induced IL-6 concentration and 30-day hospital readmission (p = 0.015), indicating that for every unit increase in IL-6, the odds of readmission increase slightly. Additionally, there was a significant interaction between the sepsis versus CINS subgroups and hospital readmission (p = 0.045), with sepsis patients showing markedly higher odds of 30-day readmission compared to CINS controls (odds ratio = 27.616). While the total number of hospital readmissions was low (four readmissions among 28 septic patients who were alive at 30 days, and 2 readmissions among 25 CINS patients who were alive at 30 days), these findings may suggest that elevated endotoxin-induced IL-6 concentrations and sepsis status are important predictors of readmission.

## 3. Discussion

Venet et al. recently identified significant immune changes by the end of the first week post-admission for severe injury, associated with an increased risk of secondary infections, and termed ‘delayed injury-acquired immunodeficiency’ [25]. In a similar, though smaller, prospective observational analysis, we delved deeper into different immune surveillance markers expressed during the first two weeks of critical injury. All the biomarkers of immunoparalysis employed in the present study significantly increased over this recovery period following severe injury. When comparing sepsis and CINS subgroups, however, we noted that temporal increases in mHLA-DR were primary driven by the CINS subset of patients, while the septic subset drove increases in ALC, LPS-induced TNF and IL-6 (whether normalized to monocyte count or not), and PMA-induced IFNγ in the overall cohort. While immunoparalysis biomarkers were not associated with 30-day mortality, stimulated IL-6 production was associated with 30-day hospital readmission in the septic subset.

In a recent, retrospective analysis, Adigbli et el. reported that early, persistent lymphopenia (defined as ALC <1.0× 10_9_/L on at least 2 days within the first 4 days of ICU admission) was associated with increased risk of death in critically ill patients with and without sepsis, with the former group demonstrating a stronger association (hazard ratios 1.89 and 1.17, respectively) [33]. Using data from patients who had ALC measured on days 1 and 7 of critical illness, we did not observe a similar association. This lack of association may be attributed to the smaller population size in our study. Mean ALC was significantly different between sepsis and CINS subgroups within the first week following enrollment, although there was no difference between groups at day 14. Surprisingly, despite lymphocytes being the predominant cellular source of IFNγ, ALC was only weakly correlated with stimulated IFNγ production over time. However, patterns of IFNγ produced in response to anti-CD3/anti-CD28 and PMA remained moderately correlated.

The cytokine response pattern to ex vivo endotoxin stimulation paralleled that of ALC. Specifically, mean TNF concentrations significantly differed between sepsis and CINS subgroups on days 1 (p < 0.001) and 7 (p = 0.001), but not on day 14. Similarly, mean IL-6 concentrations showed significant differences between subgroups on days 1 (p =0.0034) and 7 (p < 0.001), with no significant difference on day 14 (p = 0.147). Specific T lymphocyte stimulation resulted in a similar pattern in IFNγ production (p = 0.048 on day 1 and p = 0.043 on day 7, with no significant difference on day 14). Non-specific T lymphocyte stimulation with PMA also reflected this pattern (p = 0.006 on day 1, p = 0.030 on day 7, and p = 0.493 on day 14). These findings suggest that the inflammatory response is acute and pronounced during the early stages of critical illness but tends to normalize by day 14. Comparison of immunoparalysis biomarkers between healthy volunteers and critically ill patients, over a 14-day period, may shed light on any residual deficits in immune function over the study period.

Several prior studies have described mHLA-DR as a marker of monocyte deactivation or evolving immunoparalysis, especially in the context of sepsis [34,35]. Deactivated monocytes are characterized by a loss of antigen-presenting capacity and decreased reduction of their ability to produce endotoxin-induced TNF in vitro [26,36]. De Roquetaillade et al. found that a decrease or continued low expression of mHLA-DR within the first week of ICU admission was independently linked to a higher risk of subsequent infections [37]. However, mHLA-DR has several critical shortcomings that preclude its use as a biomarker in the clinical setting, including its dependence on flow cytometry and variability in measurement and analysis [38,39]. In this respect, ALC or ex vivo assays using rapid and automated cytokine measurement platforms may offer a feasible alternative for point-of-care-testing.

In the present study, we did observe that mHLA-DR significantly recovered over time, although this increase in expression was predominant in the CINS subgroup. Considering Venet et al.’s findings [25], however, it might be more appropriate and timelier to move beyond the traditional ‘sepsis’ versus ‘non-sepsis’ classifications when studying immunoparalysis in critically ill patients.

This study had several limitations that should be acknowledged. First, due to the time-consuming nature of flow cytometry, we preserved blood samples in a protein-stabilizing solution to allow for subsequent batch processing of HLA-DR quantification. While this method is established for banking human-derived samples [40-42], the freeze-thaw process may have compromised cell integrity, potentially leading to errors in HLA-DR quantification.

Second, the study’s relatively small sample size and single-center design may limit the generalizability of our findings. The small number of patients, especially in subgroup analyses, reduces the power to detect significant differences and may lead to type II errors.

Third, the study design did not allow for the assessment of the functional capacity of immune cells beyond cytokine production. Further functional assays could provide a more comprehensive understanding of the immune status of critically ill patients.

Fourth, our definition of immunoparalysis was based solely on immune parameters without considering clinical outcomes such as secondary infections. Although this approach standardizes the assessment of immunoparalysis, it may not fully capture the clinical relevance of the observed immune alterations. Despite these limitations, our findings provide valuable insights into the immune dynamics of critically ill patients with and without sepsis and highlight the need for more nuanced immune monitoring approaches.

## 4. Materials and Methods

### 4.1. Patient Cohort

Critically ill patients potentially suffering from sepsis were identified using a Modified Early Warning Scoring (MEWS) algorithm from November 2021 to March 2024. To ensure an unbiased selection process, dual, independent investigators reviewed electronically flagged patient records to identify those meeting inclusion criteria. Informed consent was obtained from patients with decision-making capacity or from legally authorized healthcare representatives for those without decision-making capacity.

Eligible participants were adults over the age of 18, within 48 hours of the onset of critical illness. Sepsis was defined according to the Sepsis-3 criteria [1]. Critical illness was defined by the need for ongoing noninvasive or invasive respiratory support and/or continuous intravenous vasopressor medications. Non-survivors were those who died within 30 days of enrollment. Patients receiving immunomodulating therapies were excluded. Measured clinical outcomes included 30-day mortality, in-hospital mortality, secondary infection rates, ICU length-of-stay and hospital readmission rates.

### 4.2. Cytokine Responses by Whole Blood Following Ex Vivo Stimulation

Fifty microliters of whole blood were diluted ten-fold in HEPES-buffered Roswell Park Memorial Institute (RPMI) 1640 media and exposed to specific stimulants as per established protocols [15,16]. Blood samples from each participant were treated under three conditions: (1) 500 pg/mL lipopolysaccharide (LPS) from Salmonella enterica strain abortus equi, (2) 500 ng/mL anti-CD3 with 2.5 μg/mL anti-CD28, or (3) 10 ng/mL phorbol 12-myristate 13-acetate (PMA) with 1 μg/mL ionomycin. After incubation for 18 hours at 37°C with 5% CO2, concentrations of IFNγ, TNF, and IL-6 were measured in triplicate using the Ella™ automated immunoassay system (Bio-Techne, Minneapolis, MN). Data were processed using Simple Plex Runner software v.3.7.2.0 (ProteinSimple) and were available within 90 minutes.

The production of TNF was normalized to the monocyte count, and IFNγ production was normalized to the lymphocyte count, based on daily complete blood counts with automated differential profiles [16,43]. Due to the varied cellular sources of IL-6, its production was normalized to the total leukocyte count [27].

### 4.3. Assessment of mHLA-DR expression

One milliliter of venous blood was collected in a tube containing sodium heparin. A proteomic stabilizer (cat# 501351689, Smart Tube Inc, Las Vegas, NV) was added to the blood prior to freezing at -80°C. At the time of mHLA-DR quantification, blood was thawed per manufacturer’s instructions. Quantibrite Anti-HLA-DR/Anti-Monocyte antibody (cat# 340827 BD Biosciences, San Diego, CA) was used to stain samples for flow cytometric quantification of mHLA-DR expression. Analysis was performed using a FACS Symphony A3 (Becton Dickson & Company, Franklin Lakes, NJ) and using Flowjo v10.8.1 (BD Biosciences).

### 4.4. Statistical Analysis

Using SAS (v9.4; SAS Institute, Cary, NC), with a significance threshold of 0.05, we summarized variables descriptively. The distribution of continuous variables was evaluated via histograms, probability plots, and normality tests. Demographic comparisons between groups utilized Chi-square and two-sample t-tests. We employed correlation specific to repeated measures taken over time to examine the relationships among various markers of immunoparalysis.

We used a linear mixed-effects model to compare cytokine levels between septic patients and CINS controls. Due to the non-normal distribution of cytokine data, log transformation was applied before modeling. Mean cytokine concentrations were then back-transformed to their original scale, and percentage change was calculated by exponentiating the slope estimates from the model, facilitating the interpretation of cytokine dynamics. For binary outcome variables including in-hospital mortality, 30-day mortality, 30-day readmission, and secondary infections, we used a binomial logistic regression model that included factors for each cytokine on day 1 and study group. Odds ratios overall and within each study group were used to quantify the magnitude of the relationship between each biomarker and outcome variable.

## 5. Conclusions

This study demonstrates that immune function in critically ill patients varies significantly over the first two weeks following the onset of sepsis. Monocyte HLA-DR expression increased mainly in non-septic patients, while ALC and ex vivo cytokine production showed significant recovery in septic patients. These findings highlight the importance of nuanced immune monitoring beyond traditional sepsis classifications. Further research is needed to refine immunoparalysis markers and improve patient outcomes.

## Author Contributions

Conceptualization, A.S.B.; methodology, A.S.B.; formal analysis, E.L.; investigation, A.S., A.S.B.; resources, A.S.B.; data curation, A.S.; writing—original draft preparation, A.S.B., A.S., P.B.; writing—review and editing, A.S.B., A.S., P.B., E.L.; visualization, A.S.B., E.L.; supervision, A.S.B.; project administration, A.S.B.; funding acquisition, A.S.B. All authors have read and agreed to the published version of the manuscript.

## Funding

This research was funded by the National Institute of General Medical Sciences, grant number R35GM150695 (ASB).

### Institutional Review Board Statement

The study was conducted in accordance with the Declaration of Helsinki and approved by the Institutional Review Board (or Ethics Committee) of the Human Studies Protection Office of the Penn State College of Medicine (protocol code #15328, approved 7/30/2020).

### Informed Consent Statement

Informed consent was obtained from all subjects involved in the study.

## Data Availability Statement

The datasets used and/or analyzed during the current study are available from the corresponding author on reasonable request.

## Acknowledgments

Penn State Critical Illness and Sepsis Research Center (CISRC).

## Conflicts of Interest

The authors declare no conflicts of interest. The funders had no role in the design of the study; in the collection, analyses, or interpretation of data; in the writing of the manuscript; or in the decision to publish the results.

## Appendix A

**Table A1.**
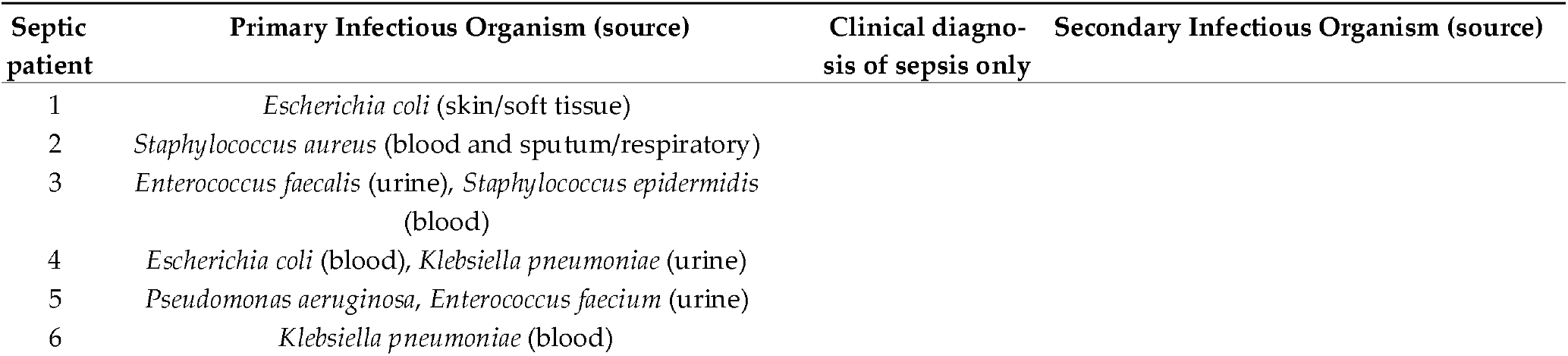
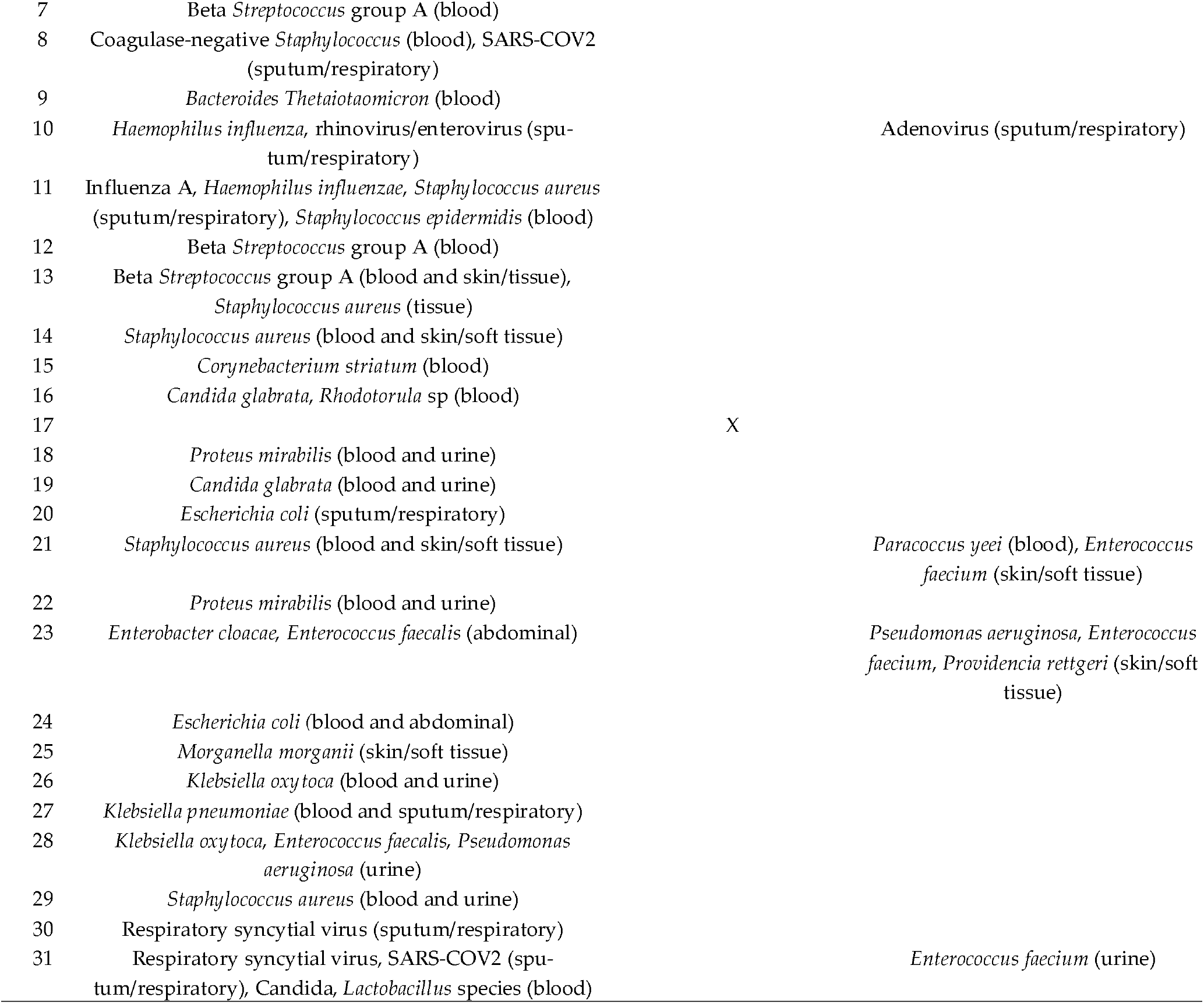
Microbial sources of sepsis.

**Table A2:**
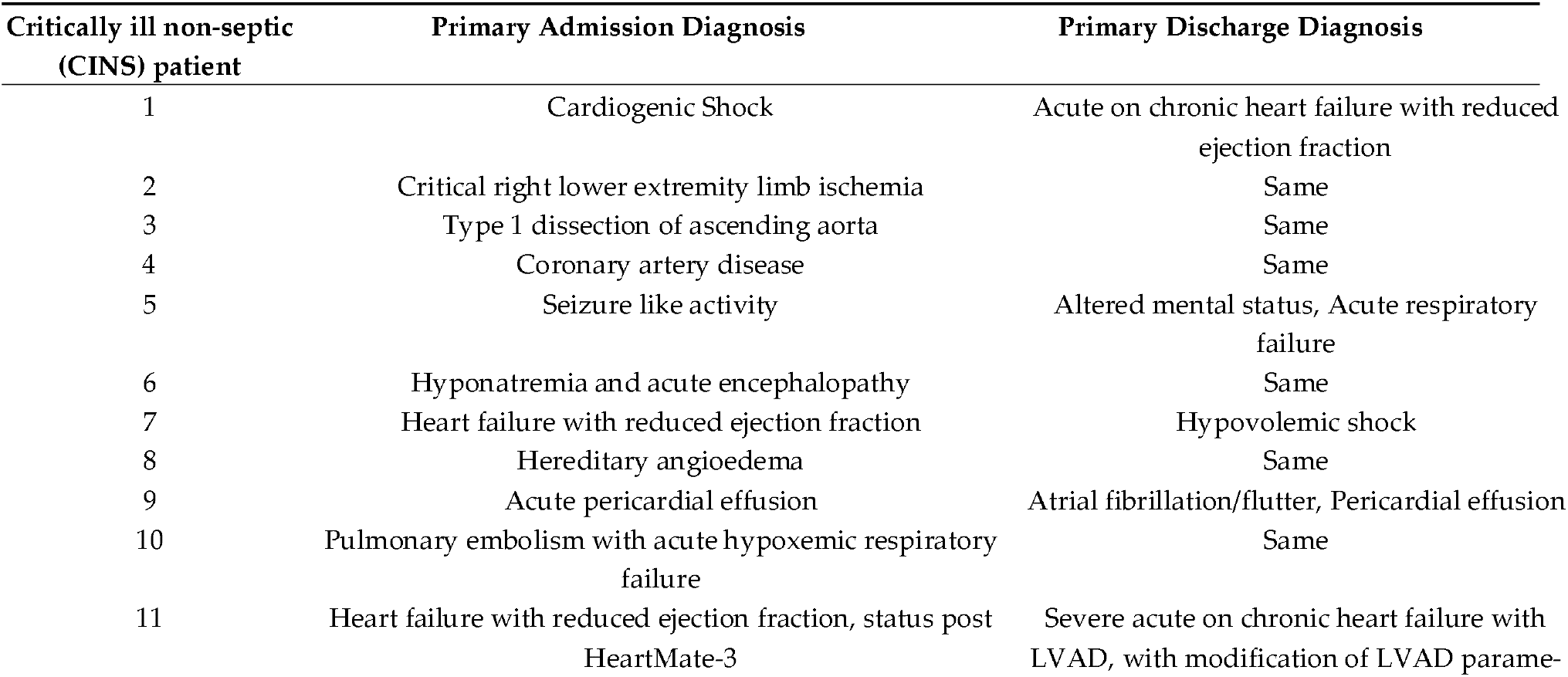
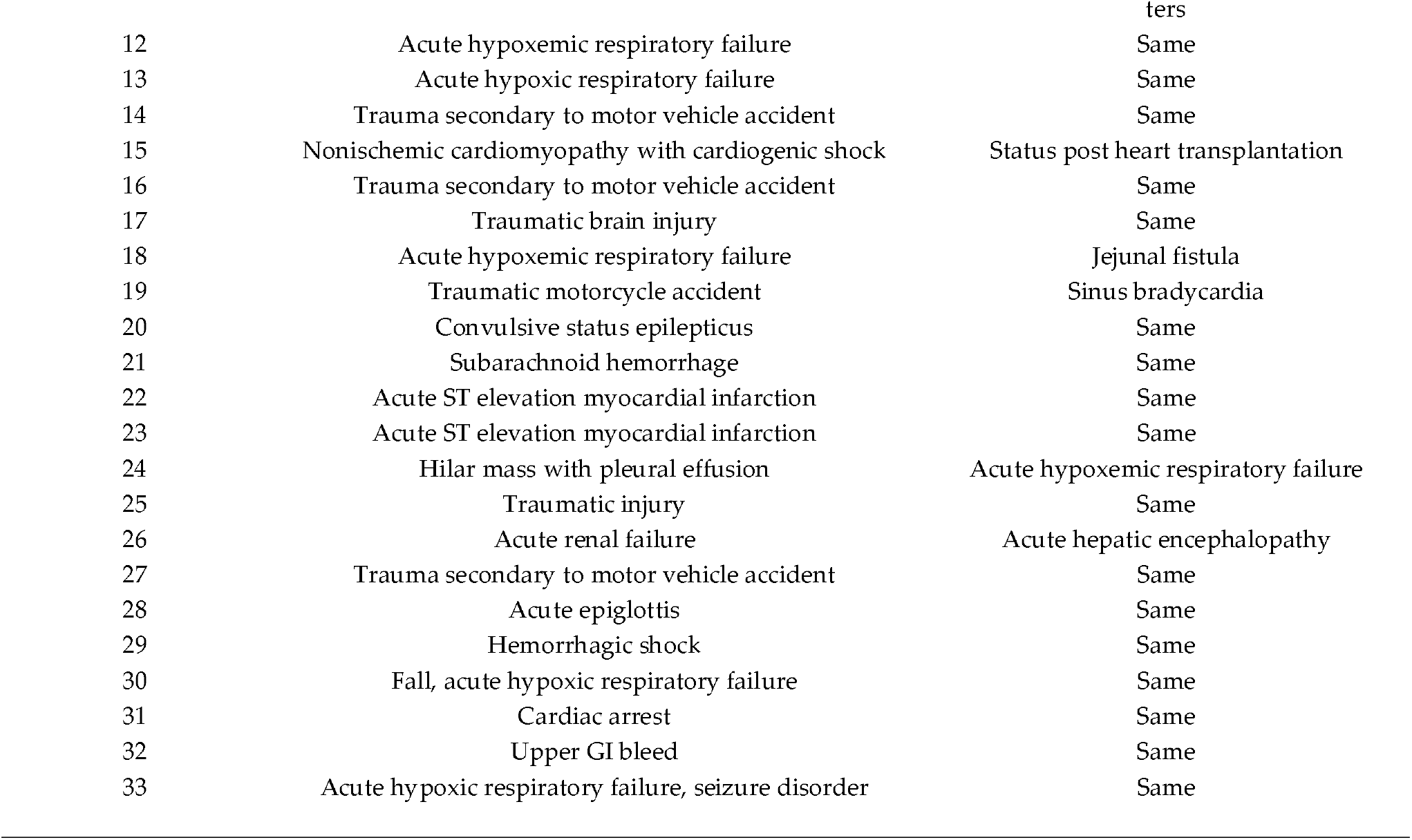
Disease etiologies of patients having non-septic, critical illness.

## Appendix B

**Figure B1.**
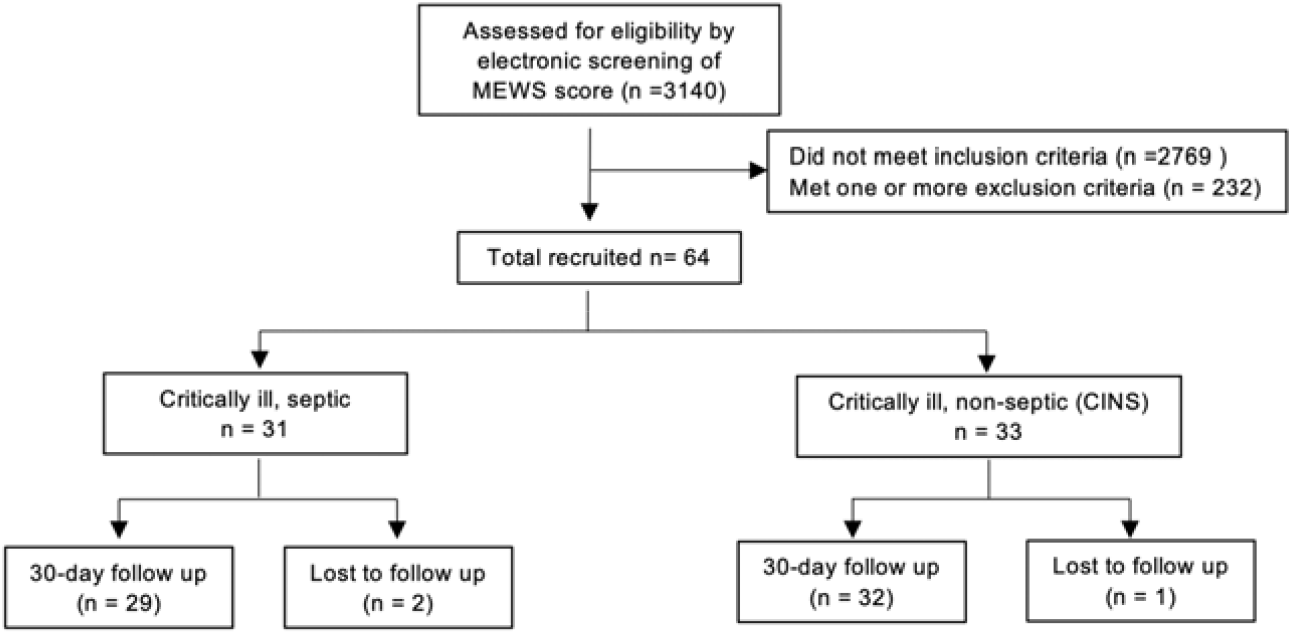
Flow diagram demonstrating the number of individuals included at each stage of the study.

## Notes

### Competing Interest Statement

The authors have declared no competing interest.

### Author Declarations

IRB (Human Subjects Protection Office) of Penn State Hershey Medical Center gave ethical approval for this work

